# Wearable tissue oximetry during standardized physiological stressors in chronic heart failure outpatients

**DOI:** 10.64898/2026.07.07.26357512

**Authors:** Thibault Roumengous, Aiden J. Chauntry, Christopher Flippen, Josephine Wallner, David A. Baran, Douglas D. Harkins

## Abstract

**Background:** Outpatient chronic heart failure (HF) assessment relies on NYHA class and distance-based testing that can obscure physiological heterogeneity. Near-infrared spectroscopy (NIRS) enables tissue oxygenation phenotyping but is underexplored during standardized stressors in outpatient HF. We tested whether wearable NIRS-derived oxygenation kinetics during a vascular occlusion test (VOT) and six-minute walk test (6MWT) differ across NYHA classes.

**Methods:** In this prospective, single-center pilot study, 44 chronic HF outpatients (mean age 70.9 ± 8.7 years, 75% male; NYHA I [n=19], II [n=12], III [n=13]) were monitored with a novel wearable NIRS device (NIRSense Envello Core) during a VOT and 6MWT. Primary endpoints were the post-occlusion net area under the curve (net AUC; VOT) and post-walk recovery net AUC (modified 6MWT). Secondary endpoints included the exertional tissue oxygenation (Oxy) nadir, VOT reperfusion kinetics, gait metrics, and tolerability.

**Results:** Despite NYHA I and II walking identical median distances (420 m), post-walk recovery net AUC was lower in NYHA II (−16.3 a.u.×s) and III (−12.8 a.u.×s) than NYHA I (46.1 a.u.×s; p=0.004). The exertional Oxy nadir did not differ (p=0.722), but NYHA III walked 27% and 38% slower than NYHA II and I (p<0.001). NYHA II had higher VOT net AUC (134.2 a.u.×s) than NYHA I (71.9; p=0.018) and III (61.1; p=0.011). Post-walk recovery net AUC correlated with gait velocity (r_s_=0.44) and distance (r_s_=0.39; both p<0.05). VOT net AUC did not correlate with functional metrics, but resting reperfusion kinetics correlated with 6MWT performance (r_s_=0.41–0.46, p<0.05). The sensor was well tolerated.

**Conclusions:** Wearable NIRS-derived recovery kinetics differentiated NYHA I from NYHA II despite these classes walking identical median distances. Coupled with distinct resting VOT hyperemic differences, these preliminary findings indicate wearable NIRS may capture physiological heterogeneity in outpatient HF not reflected by NYHA class and standard functional metrics.

## Introduction

Chronic heart failure (HF) is characterized by cardiac dysfunction and abnormalities in microvascular and skeletal muscle oxygenation that contribute to impaired local tissue perfusion and exertional intolerance^1^. Management of chronic HF remains heavily reliant on the New York Heart Association (NYHA) functional classification^2–4^, which is subjective and lacks the precision to capture underlying microvascular heterogeneity^3^. When functional capacity is evaluated clinically in chronic HF, the 6-minute walk test (6MWT) is a standard submaximal assessment, but behavioral self-pacing may obscure underlying physiological heterogeneity^5^ and traditional outcomes like total distance often fail to differentiate early disease stages^6^. This can limit the ability to detect clinically relevant physiological changes before overt deterioration occurs. Measuring tissue-level oxygenation responses during the 6MWT and other stressors, such as a vascular occlusion test, is not routine in outpatient HF care^2–4^. However, this may provide important clinical information that is not captured by symptom-based clinical stratification or traditional 6MWT endpoints alone^1,4^, thereby allowing for more objective disease severity stratification.

Near-infrared spectroscopy (NIRS) offers a noninvasive optical approach for continuously monitoring oxygenation profiles during stressors^7^. A VOT applies a controlled ischemia-reperfusion perturbation from which microvascular reactivity and recovery can be quantified^7^. The VOT elicits a characteristic tissue oxygenation trajectory (rapid desaturation during occlusion followed by a reactive hyperemic overshoot upon cuff release) that yields time-resolved indices of microvascular reserve^8,9^. In critical illness, NIRS-derived VOT reperfusion metrics carry prognostic value, such that blunted reperfusion slopes predict organ dysfunction and mortality^10,11^, and NIRS-VOT reactive hyperemia is disrupted in individuals at high cardiovascular risk^12^. The 6MWT assesses metabolic oxygen demand during functional effort and post-exercise recovery^5,13^, and together with the VOT can capture distinct and complementary aspects of tissue hemodynamics.

NIRS has been used in HF to characterize muscle deoxygenation during exercise^14–16^ and resting muscle oxygen consumption during arterial occlusion^17^. However, translation to HF outpatient workflows has been limited^18^, likely reflecting prior device constraints, including tethered form factors, limited motion tolerance, and operational complexity incompatible with ambulatory clinical workflows^7,18–20^. While miniaturized wearable clinical NIRS platforms may overcome these barriers and complement functional assessments and NYHA classification, their utility for continuously monitoring tissue oxygenation responses during functional and VOT testing in HF outpatients remains unstudied.

This prospective proof-of-concept study aimed to evaluate a novel wearable NIRS-based tissue oximeter (NIRSense Envello Core) during a 6MWT and VOT in outpatients with chronic HF (NYHA I–III). The primary objective was to test for differences in the net baseline-referenced area under the oxygenation–time curve (net AUC; a.u. × s) across NYHA functional classes I–III, computed over the 3-minute post-walk recovery window (6MWT) and the 3-minute post-cuff-release window (VOT). We hypothesized that more symptomatic patients would demonstrate altered tissue oxygenation kinetics, including attenuated recovery or reperfusion responses. Exploratory objectives were to examine (1) additional VOT and 6MWT oxygenation features by NYHA class, (2) associations between VOT endpoints and 6MWT metrics, as well as 6MWT functional and NIRS metrics, (3) short-interval VOT test-retest reliability and trial-order effects, and (4) patient acceptability and tolerability of the wearable sensor. We hypothesized that secondary oxygenation-derived features would show similar NYHA differences, that VOT reperfusion features would correlate with 6MWT functional metrics, that 6MWT functional endpoints would be associated with 6MWT NIRS endpoints, that primary VOT endpoints would demonstrate acceptable short-interval repeatability without systematic trial-order effects, and that the wearable form factor would yield high patient acceptability.

## Methods

### Study Design and Setting

This prospective, single-center pilot observational study was conducted in an outpatient cardiology practice (Jackson Heart Clinic, Jackson, MS, USA) and approved by the WIRB–Copernicus Group Institutional Review Board (IRB #20233808). All participants provided written informed consent. During a routine clinical visit, participants were instrumented with two wearable sensors (NIRSense Envello Core) and completed a VOT and 6MWT, separated by 10 minutes. All testing was performed in designated clinical spaces with consistent ambient lighting and temperature. Given the pilot nature of the study, no formal power calculation was conducted.

### Participants

A pragmatic sample of 44 adults was recruited from the existing patient population via clinic-based approaches based on feasibility within the clinical workflow. Inclusion criteria were age 18–90 years, heart failure with NYHA functional class I–III, body mass index 18–40 kg/m², body weight >40 kg, and the ability to ambulate independently for the 6MWT. Exclusion criteria were self-reported allergy to medical-grade adhesives, pregnancy, history of sickle cell trait, anemia, or thalassemia, and NYHA class IV. The collection of demographic and clinical data, including Fitzpatrick skin phototype, is detailed in Supplementary Text 1. NYHA functional class (I–III) was derived from each participant’s most recent medical record and physician-adjudicated on the day of enrollment.

### Standardized physiological stressors

#### Vascular occlusion test (VOT)

Each participant completed two seated VOT trials with the arm resting on a pillow at heart level. The second VOT trial was performed only after StO2 returned to the pre-VOT baseline value (within 1% for at least 30 s) and a minimum of 10 minutes of recovery had elapsed. The NIRSense Envello Core was secured distal to a brachial cuff on the ipsilateral forearm. After a 10-minute baseline, the cuff was rapidly inflated to 50 mmHg above systolic blood pressure and maintained until StO2 reached 40% or for a maximum of 3 minutes, whichever occurred first^9^, then rapidly deflated. For participants with two analyzable trials, endpoints were calculated for each trial and averaged to yield participant-level values; when only one analyzable trial was available, that value was used. Further details are described in Supplementary Text 1.

#### Six-minute walk test (6MWT)

A modified 6MWT protocol adapted from ATS procedures was employed^21^. The timer was paused during rest breaks and resumed upon recommencement of ambulation to ensure all participants completed a cumulative 360s of active locomotion and equalize the metabolic stimulus duration for NYHA comparisons. Accordingly, distance outcomes from this modified protocol are not directly comparable with standard ATS 6MWT norms.

### NIRSense Envello Core

#### Device details

The NIRSense Envello Core (NIRSense, Richmond, VA, USA; Figure 1) is an investigational wearable optical monitor (75 × 35 × 13 mm; 41 g) that acquires tissue oxygenation by continuous-wave NIRS (modified Beer–Lambert principle) and is capable of photoplethysmography and 3-axis accelerometry. Two sensors were applied per participant: longitudinally over the extensor carpi radialis of the dominant forearm (VOT analyses) and vertically over the mid-sternum (6MWT analyses); the sternal site was chosen to reduce locomotor-motion artifact and reflects a mixed systemic-regional signal. The NIRSense Envello Core is currently undergoing reference-oximetry validation (ClinicalTrials.gov, NCT07032831). Full specifications (Supplementary Table S1) and sensor preparation details are provided in Supplementary Material.

**Figure 1.**
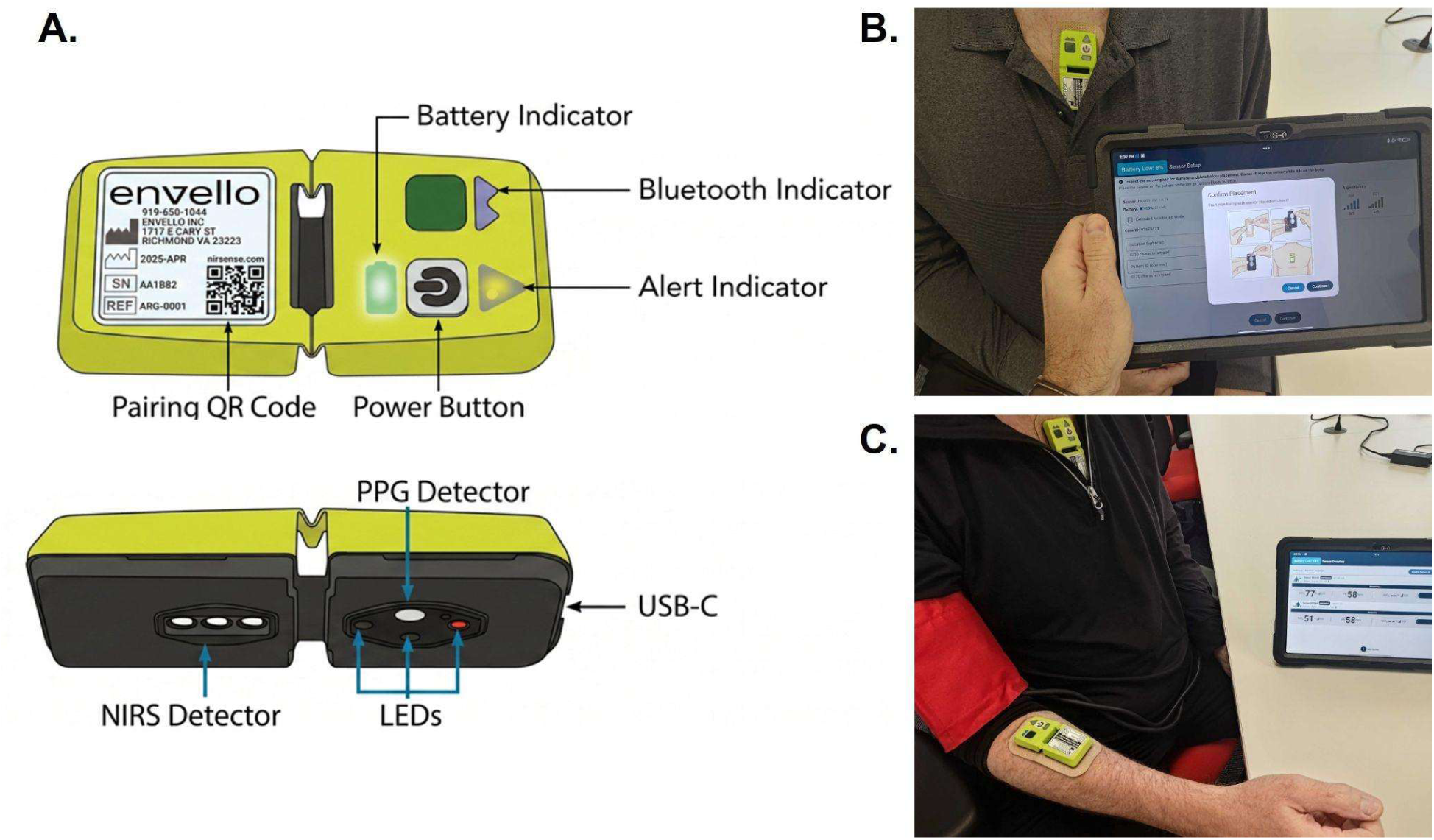
NIRSense Envello Core wearable optical sensor and participant setup. **(A)** External views of the sensor with labeled components. The front face shows the pairing quick-response (QR) code, the battery, Bluetooth, and alert status indicators, and the power button; the tissue-facing surface shows the near-infrared spectroscopy (NIRS) detector, the photoplethysmography (PPG) detector, the light-emitting diodes (LEDs), and the USB-C charging port. **(B)** Sternal sensor placement used for six-minute walk test (6MWT) analyses, with the sensor positioned vertically over the mid-sternum; the paired Android tablet displays the NIRSense application placement-confirmation screen. **(C)** Forearm sensor placement used for vascular occlusion test (VOT) analyses, with the sensor secured over the dominant forearm distal to a brachial blood-pressure cuff; the tablet displays real-time signal acquisition. Detailed device dimensions, optical and sampling specifications are provided in Supplementary Table S1.

#### Signal processing

Oxygenation features were derived from the 35-mm source-detector channel. Continuous signals were low-pass filtered, downsampled to 1 Hz, smoothed, and baseline-referenced (zero-centered) to each stressor; trials with severe motion, ambient light, or decoupling artifacts were excluded. Because all endpoints were baseline-referenced within-subject signals (arbitrary units) rather than absolute StO2, they depend on the device resolving directional and kinetic change rather than absolute calibration^22,23^. Full processing parameters are provided in Supplementary Text 1.

### Primary and secondary landmarks and endpoints

All endpoints were derived from the zero-centered relative tissue oxygenation (Oxy) signals. The primary VOT endpoint (Figure 2) was the post-occlusion net baseline-referenced area under the oxygenation–time curve (post-occlusion net AUC; a.u. × s), integrated (trapezoidal rule) from cuff release over a 3-minute recovery window. The primary 6MWT endpoint (Figure 3) was the post-walk recovery net AUC, integrated over a 3-minute window immediately following cessation of ambulation. Secondary VOT endpoints were the maximum reperfusion slope and the reactivity ratio, as well as ischemic-stimulus metrics (occlusion duration, ischemic nadir, maximum deoxygenation slope). Secondary 6MWT endpoints were the exertional Oxy nadir, total distance, active gait velocity, rest-break frequency, and total elapsed time. Detailed landmark definitions are provided in Supplementary Text 1.

**Figure 2.**
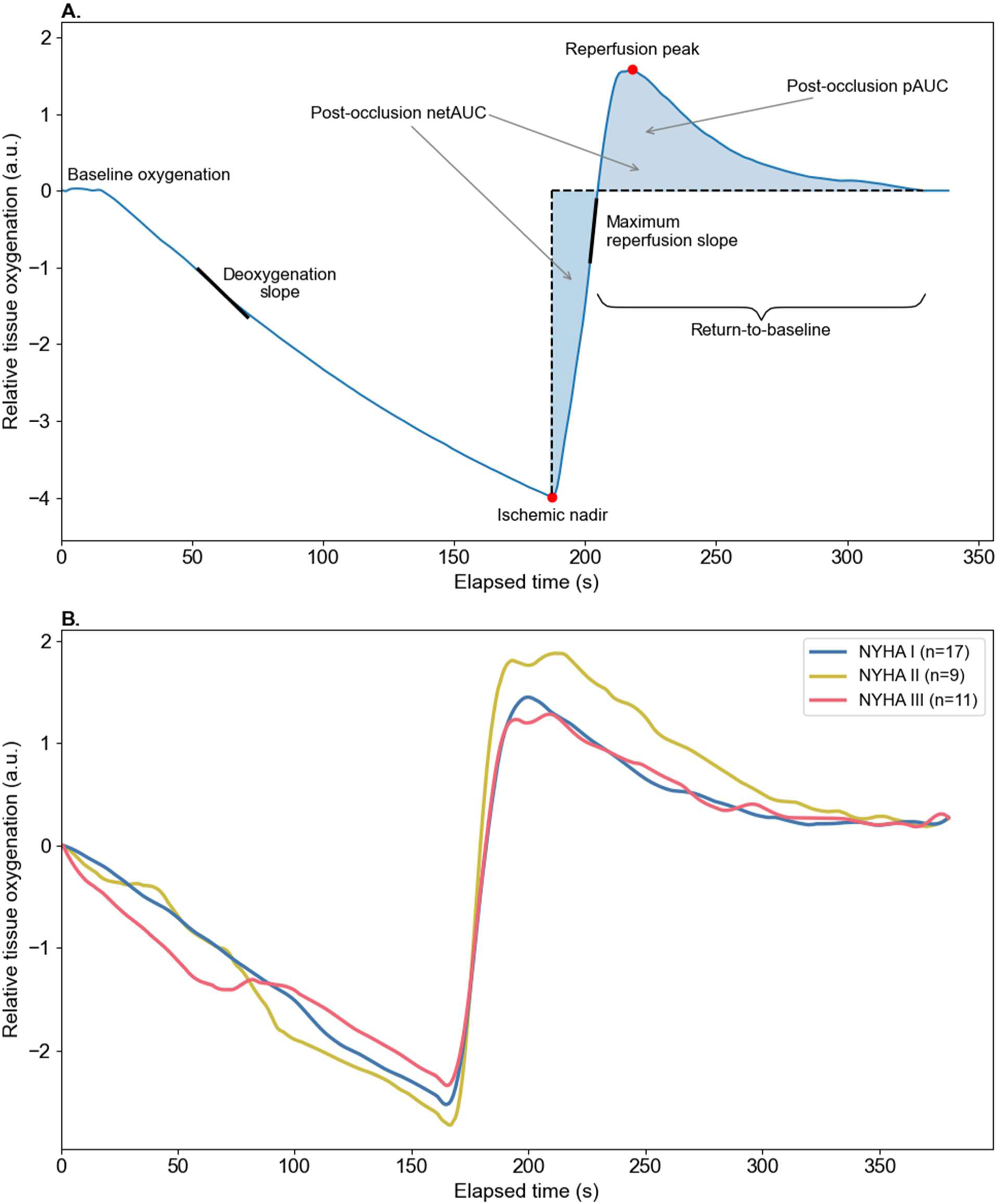
Vascular occlusion test (VOT) feature landmarks and New York Heart Association functional class comparison. **(A)** Representative zero-centered relative tissue oxygenation (Oxy) trace from a single participant during the VOT protocol. Annotated landmarks include the resting baseline, deoxygenation (Deoxy) slope, ischemic nadir, maximum reperfusion (Reoxy) slope, and reperfusion peak. The post-occlusion net area under the curve (net AUC) is represented by the shaded region. **(B)** Group-level Oxy time series stratified by NYHA functional class (I: n=17; II: n=9; III: n=11), with all traces aligned to the initiation of cuff inflation (time = 0), which was recorded during each VOT. Lines represent smoothed group medians; shaded interquartile range (IQR) was omitted from the main figure to enhance visual clarity of divergent kinetic trajectories across NYHA classes (see Supplementary Figure S1 for full data distribution)

**Figure 3.**
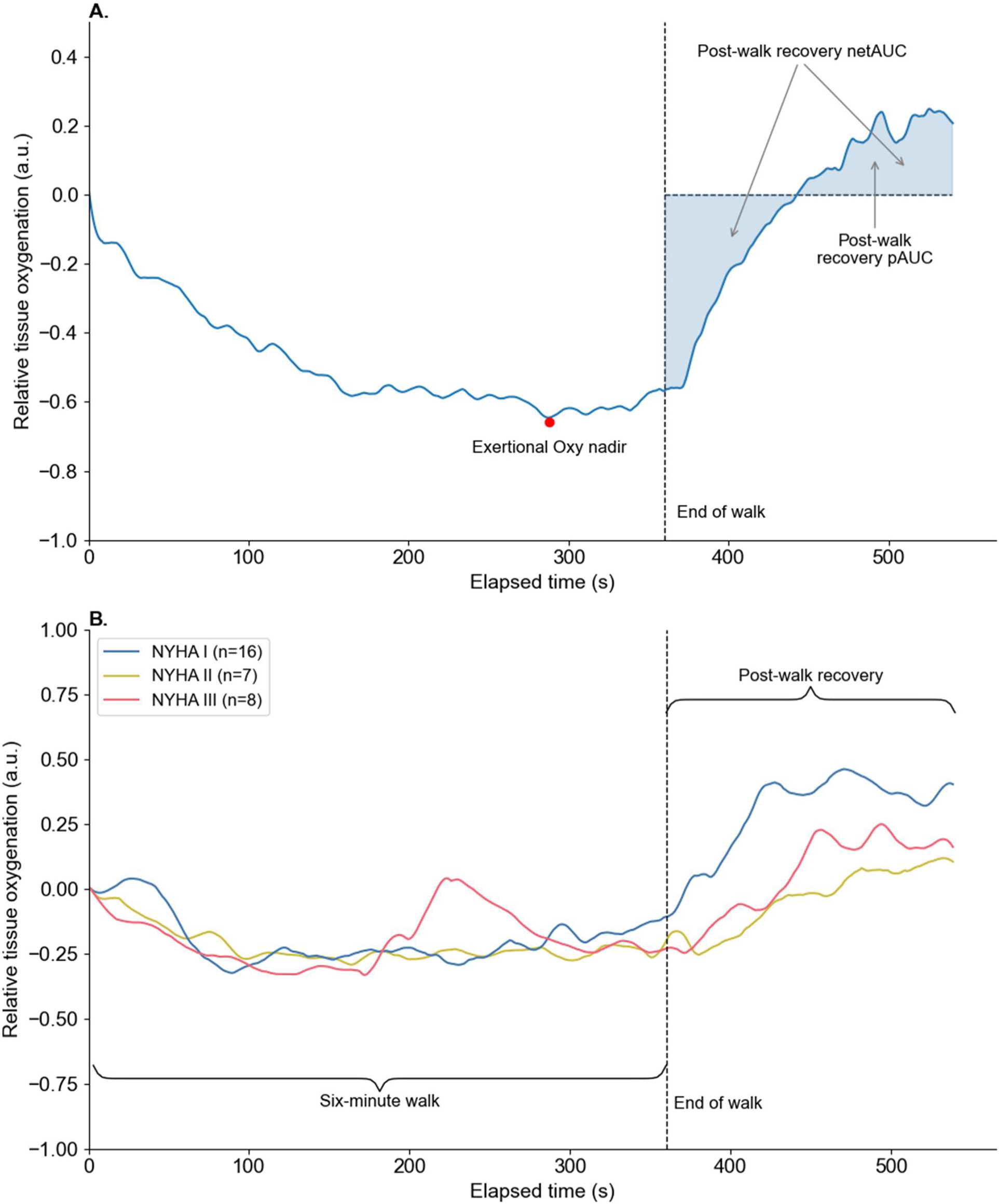
Sternal tissue oxygenation kinetics during and after the 6-minute walk test. Note. **(A)** Representative relative tissue oxygenation (Oxy) trace from a single participant illustrating primary and secondary ambulatory endpoints. The shaded region denotes the post-walk recovery net AUC (net AUC), integrated over a 3-minute window following exercise cessation at 360s. **(B)** Group-level kinetic trajectories across NYHA classes (I: n=16; II: n=7; III: n=8). Lines represent smoothed group medians; shaded interquartile range (IQR) was omitted from the main figure to enhance visual clarity of divergent kinetic trajectories across NYHA classes (see Supplementary Figure S2 for full data distribution). a.u. = arbitrary units; NYHA = New York Heart Association.

### Patient Acceptability Survey Questions

Participants completed a study-specific questionnaire evaluating device and adhesive comfort, willingness for extended wear, and anticipated ease of self-application (instrument detailed in Supplementary Text 1).

### Statistical Analysis

Analyses were performed in Python (version 3.12.13; pandas, scikit-learn, NumPy, scikit-posthocs, SciPy, pingouin). Statistical significance was two-tailed p < 0.05. Given the sample size, interpretation should emphasize effect sizes and the distribution of medians rather than p-values alone. Normality was assessed with the Shapiro-Wilk test and histograms. Continuous variables were reported as mean ± SD or median [IQR] and categorical variables as n (%), in total and across NYHA class. Between-group differences across NYHA classes (I–III) were tested with Kruskal-Wallis tests and epsilon-squared (ε²) effect sizes. A single primary endpoint was pre-specified for each stressor (post-occlusion net AUC for the VOT, post-walk recovery net AUC for the 6MWT); all other endpoints were secondary, unadjusted for multiple comparisons, and interpreted as hypothesis-generating. Significant Kruskal-Wallis results were followed by Dunn’s post-hoc tests with Holm-Bonferroni adjustment and Cliff’s delta (δ); non-significant endpoints were reported descriptively. Relationships between VOT and 6MWT metrics, and between 6MWT functional and NIRS metrics, were evaluated with Spearman’s rank-order correlations. Complete-case analyses were used given the pilot sample size and endpoint-specific signal loss^24^. Test-retest reliability, absolute variability, subgroup-representativeness comparisons, and sensitivity analyses are detailed in the Supplementary Text 1.

## Results

### Participant characteristics

Forty-four predominantly HFrEF patients were included (NYHA I: n=19; II: n=12; III: n=13; Table 1). The cohort was older (mean age 70.9 ± 8.7 years), predominantly male (75%), and included broad Fitzpatrick skin-type representation (types V–VI, 25%) and both ischemic (41%) and non-ischemic (59%) etiologies. Guideline-directed medical therapy use was high across classes. Beta-blocker prescription was non-monotonic (lowest in NYHA II, 75%, versus 95% and 92% in NYHA I and III), whereas loop/thiazide diuretic and SGLT2 inhibitor use rose with class, and mineralocorticoid receptor antagonist use was also non-monotonic (NYHA I, 58%; II, 25%; III, 69%). Full characteristics are provided in Table 1.

**Table 1.**
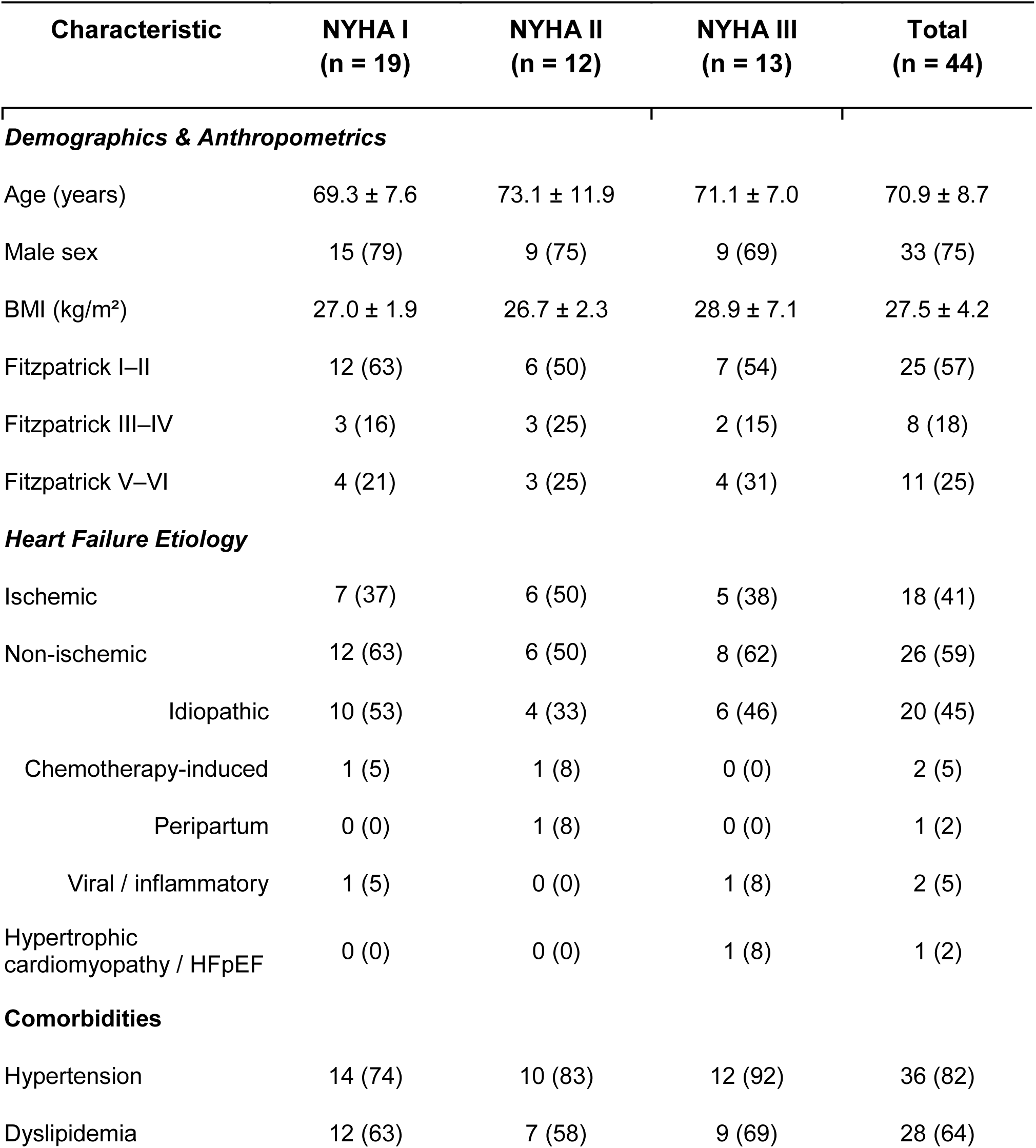
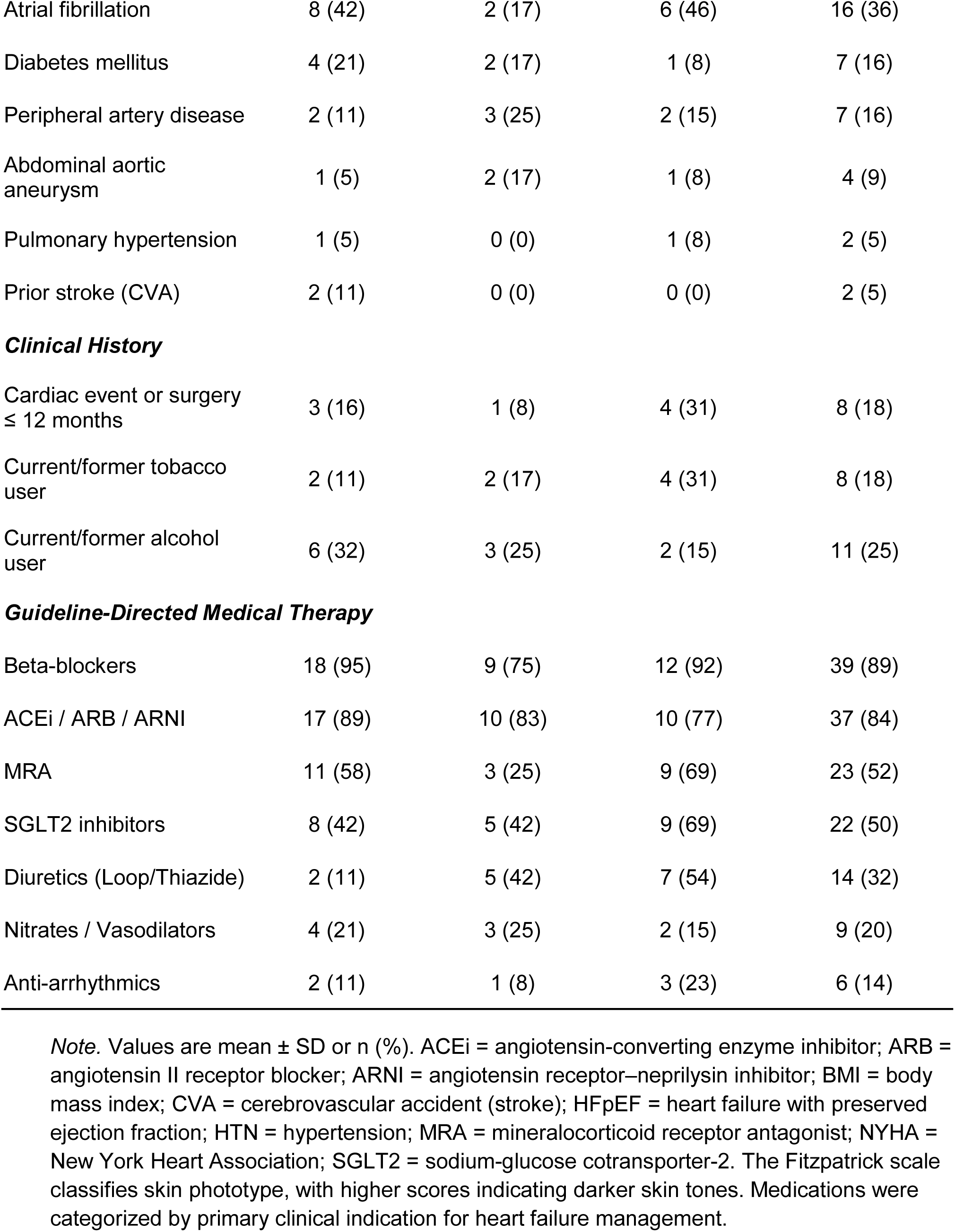
Participant characteristics stratified by New York Heart Association functional class.

| Characteristic | NYHA I<br>(n = 19) | NYHA II<br>(n = 12) | NYHA III<br>(n = 13) | Total<br>(n = 44) |
| --- | --- | --- | --- | --- |
| ***Demographics & Anthropometrics*** |  |  |  |  |
| Age (years) | 69.3 ± 7.6 | 73.1 ± 11.9 | 71.1 ± 7.0 | 70.9 ± 8.7 |
| Male sex | 15 (79) | 9 (75) | 9 (69) | 33 (75) |
| BMI (kg/m<sup>2</sup>) | 27.0 ± 1.9 | 26.7 ± 2.3 | 28.9 ± 7.1 | 27.5 ± 4.2 |
| Fitzpatrick I–II | 12 (63) | 6 (50) | 7 (54) | 25 (57) |
| Fitzpatrick III–IV | 3 (16) | 3 (25) | 2 (15) | 8 (18) |
| Fitzpatrick V–VI | 4 (21) | 3 (25) | 4 (31) | 11 (25) |
| ***Heart Failure Etiology*** |  |  |  |  |
| Ischemic | 7 (37) | 6 (50) | 5 (38) | 18 (41) |
| Non-ischemic | 12 (63) | 6 (50) | 8 (62) | 26 (59) |
| Idiopathic | 10 (53) | 4 (33) | 6 (46) | 20 (45) |
| Chemotherapy-induced | 1 (5) | 1 (8) | 0 (0) | 2 (5) |
| Peripartum | 0 (0) | 1 (8) | 0 (0) | 1 (2) |
| Viral / inflammatory | 1 (5) | 0 (0) | 1 (8) | 2 (5) |
| Hypertrophic cardiomyopathy / HFpEF | 0 (0) | 0 (0) | 1 (8) | 1 (2) |
| ***Comorbidities*** |  |  |  |  |
| Hypertension | 14 (74) | 10 (83) | 12 (92) | 36 (82) |
| Dyslipidemia | 12 (63) | 7 (58) | 9 (69) | 28 (64) |
| Atrial fibrillation | 8 (42) | 2 (17) | 6 (46) | 16 (36) |
| Diabetes mellitus | 4 (21) | 2 (17) | 1 (8) | 7 (16) |
| Peripheral artery disease | 2 (11) | 3 (25) | 2 (15) | 7 (16) |
| Abdominal aortic aneurysm | 1 (5) | 2 (17) | 1 (8) | 4 (9) |
| Pulmonary hypertension | 1 (5) | 0 (0) | 1 (8) | 2 (5) |
| Prior stroke (CVA) | 2 (11) | 0 (0) | 0 (0) | 2 (5) |
| ***Clinical History*** |  |  |  |  |
| Cardiac event or surgery<br>≤ 12 months | 3 (16) | 1 (8) | 4 (31) | 8 (18) |
| Current/former tobacco<br>user | 2 (11) | 2 (17) | 4 (31) | 8 (18) |
| Current/former alcohol<br>user | 6 (32) | 3 (25) | 2 (15) | 11 (25) |
| ***Guideline-Directed Medical Therapy*** |  |  |  |  |
| Beta-blockers | 18 (95) | 9 (75) | 12 (92) | 39 (89) |
| ACEi / ARB / ARNI | 17 (89) | 10 (83) | 10 (77) | 37 (84) |
| MRA | 11 (58) | 3 (25) | 9 (69) | 23 (52) |
| SGLT2 inhibitors | 8 (42) | 5 (42) | 9 (69) | 22 (50) |
| Diuretics (Loop/Thiazide) | 2 (11) | 5 (42) | 7 (54) | 14 (32) |
| Nitrates / Vasodilators | 4 (21) | 3 (25) | 2 (15) | 9 (20) |
| Anti-arrhythmics | 2 (11) | 1 (8) | 3 (23) | 6 (14) |

*Note.* Values are mean ± SD or n (%). ACEi = angiotensin-converting enzyme inhibitor; ARB = angiotensin II receptor blocker; ARNI = angiotensin receptor–neprilysin inhibitor; BMI = body mass index; CVA = cerebrovascular accident (stroke); HFpEF = heart failure with preserved ejection fraction; HTN = hypertension; MRA = mineralocorticoid receptor antagonist; NYHA = New York Heart Association; SGLT2 = sodium-glucose cotransporter-2. The Fitzpatrick scale classifies skin phototype, with higher scores indicating darker skin tones. Medications were categorized by primary clinical indication for heart failure management.

### Signal yield and quality control

For the VOT, 7 participants (16%) were excluded (loss of signal integrity, n=4; severe motion artifact, n=3), yielding 37 analyzable datasets (NYHA I: 17; II: 9; III: 11). For the 6MWT, 4 (9%) were excluded for data-logging errors or software instability, yielding 40 datasets; the post-walk recovery net AUC sample was further reduced to 31 (NYHA I: 16; II: 7; III: 8) owing to insufficient post-exercise recording or motion artifact during the recovery transition. Analyzable subgroups did not differ from the enrolled cohort on any measured baseline variable (all p > 0.29).

#### Vascular occlusion test reliability and trial order

Among participants with two analyzable VOT trials (n=24), test-retest reliability was acceptable to excellent (post-occlusion net AUC ICC 3,k = 0.75; maximum reperfusion slope 0.88; reactivity ratio 0.64), with no meaningful trial-order effects except a minor increase in the maximum reperfusion slope on the second trial (p = 0.037; Supplementary Table S2).

### Primary analyses

#### Post-occlusion net AUC across New York Heart Association functional classification

Post-occlusion net AUC differed across NYHA classes (H = 9.65, p = 0.008, ε² = 0.22; Table 2, Figure 4). NYHA II showed the highest values (median 134.2 a.u.×s), significantly greater than both NYHA I (71.9) and NYHA III (61.1), with no difference between NYHA I and III (Table 3). This non-monotonic pattern was unchanged in sensitivity analyses using the zero-clipped reactive-hyperemia pAUC and after outlier exclusion.

**Figure 4.**
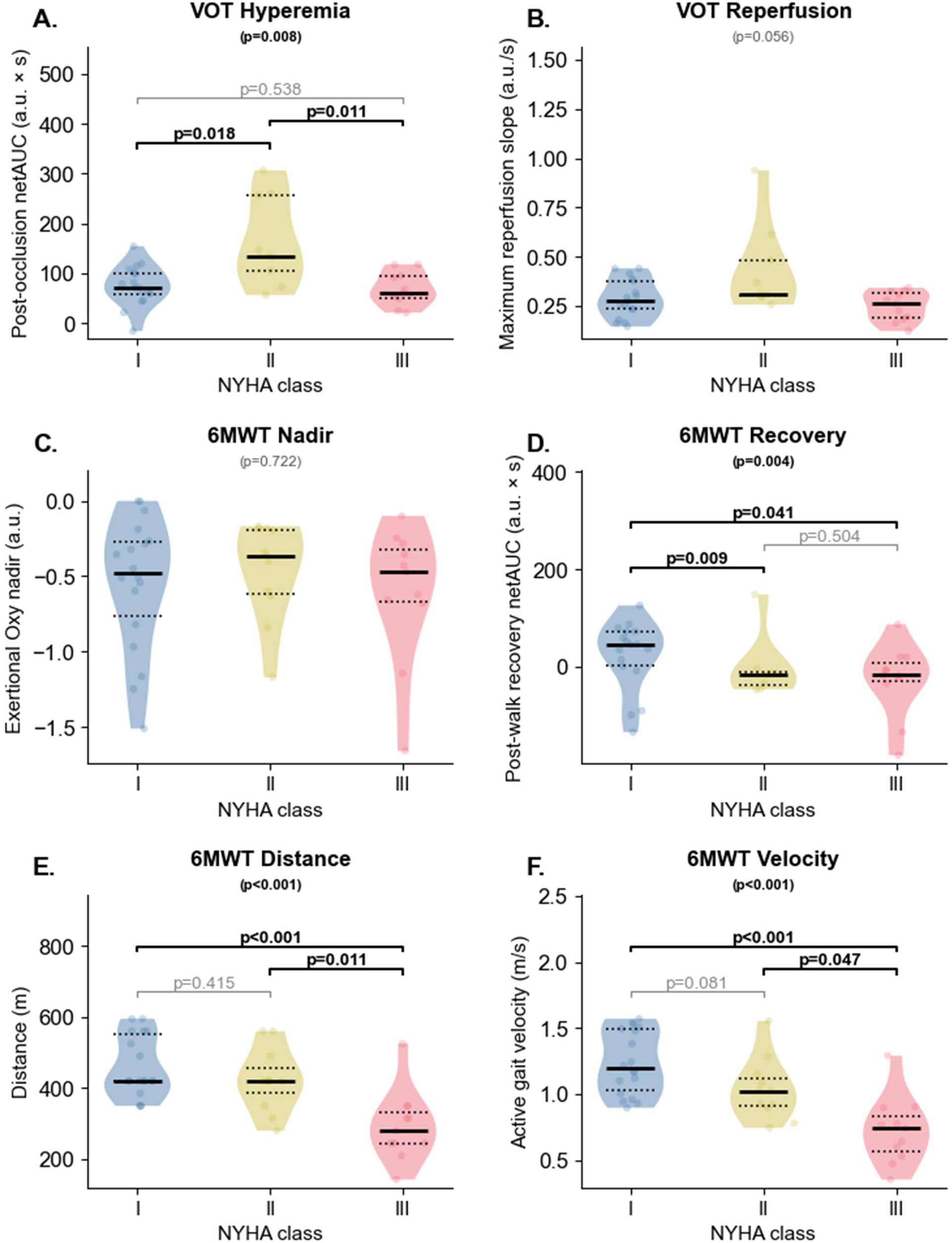
Physiological and functional endpoints across New York Heart Association (NYHA) classes. (A) Post-occlusion net area under the curve (net AUC) following resting vascular occlusion (primary VOT endpoint). (B) Maximum reperfusion slope following resting vascular occlusion. (C) Exertional tissue oxygenation nadir during the active 6-minute walk test (6MWT). (D) Post-walk recovery net AUC during the 3-minute window following the 6MWT (primary 6MWT endpoint). (E) Total distance covered during the modified 6MWT protocol. (F) Active gait velocity calculated during the 6MWT. Analyzable sample sizes vary by endpoint: VOT (n=37; NYHA I: 17, II: 9, III: 11), 6MWT primary ambulatory endpoints (n=40; NYHA I: 18, II: 11, III: 11), and 6MWT recovery net AUC (n=31; NYHA I: 16, II: 7, III: 8). Violin plots illustrate the kernel density distribution of the data. Within each violin, the solid black horizontal line denotes the median, and the upper and lower dotted lines represent the 25th and 75th percentiles (interquartile range), respectively. Semi-transparent circles represent individual patient data points. Floating p-values centered at the top of each panel denote global significance (unadjusted) derived from Kruskal-Wallis tests. Bracketed p-values denote pairwise exploratory comparisons evaluated via Dunn’s post-hoc tests with Holm-Bonferroni adjustment; significant pairwise contrasts are bolded, while non-significant contrasts are displayed unbolded in grey. a.u. = arbitrary units. Distance reflects the total distance covered during the modified 6MWT protocol.

**Table 2.** Physiological and functional kinetics during the vascular occlusion and six-minute walk tests stratified by NYHA class.

| Endpoint | NYHA I<br>Median [IQR] | NYHA II<br>Median [IQR] | NYHA III<br>Median [IQR] | p | $\epsilon^2$ |
| --- | --- | --- | --- | --- | --- |
| ***Vascular Occlusion Test (VOT) Kinetics*** |  |  |  |  |  |
| Post-occlusion net<br>AUC (a.u. × s) * | 71.9 [59.2, 101.2] | 134.2 [105.2, 256.5] | 61.1 [51.5, 93.5] | **0.008** | 0.22 |
| Max reperfusion slope<br>(a.u./s) | 0.275 [0.234, 0.377] | 0.313 [0.301, 0.483] | 0.263 [0.189, 0.315] | 0.056 | 0.11 |
| Reactivity ratio (a.u) | 7.43 [4.67, 11.02] | 11.09 [5.69, 12.67] | 8.80 [4.03, 10.02] | 0.338 | <0.01 |
| ***Six-Minute Walk Test (6MWT) Kinetics*** |  |  |  |  |  |
| Post-walk recovery net<br>AUC (a.u. × s) * | 46.1 [11.1, 72.5] | -16.3 [-39.5, -13.5] | -12.8 [-18.7, 0.72] | **0.004** | 0.32 |
| Distance during<br>modified 6MWT<br>protocol (m) | 420 [420, 551] | 420 [385, 455] | 280 [245, 333] | **<0.001** | 0.37 |
| Active gait velocity<br>(m/s) | 1.19 [1.03, 1.49] | 1.02 [0.91, 1.12] | 0.74 [0.56, 0.84] | **<0.001** | 0.44 |
| Exertional Oxy nadir<br>(a.u.) | -0.48 [-0.76, -0.27] | -0.36 [-0.61, -0.19] | -0.47 [-0.67, -0.32] | 0.722 | <0.01 |

**Note.** \* denotes primary endpoints. Analyzable sample sizes vary by endpoint: VOT (n=37; NYHA I: 17, II: 9, III: 11), 6MWT primary ambulatory endpoints (n=40; NYHA I: 18, II: 11, III: 11), and 6MWT recovery net AUC (n=31; NYHA I: 16, II: 7, III: 8). Data are presented as median [interquartile range]. Global variance across NYHA functional classes was evaluated using the Kruskal-Wallis test; corresponding p-values and epsilon-squared ( $\epsilon^2$ ) effect sizes are reported. 6MWT = six-minute walk test; a.u. = arbitrary units; NYHA = New York Heart Association; Oxy = relative tissue oxygenation; AUC = area under the curve; VOT = vascular occlusion test. Bold text indicates statistical significance ( $p < 0.05$ ). Distance reflects the total distance covered during the modified 6MWT protocol.

**Table 3.**
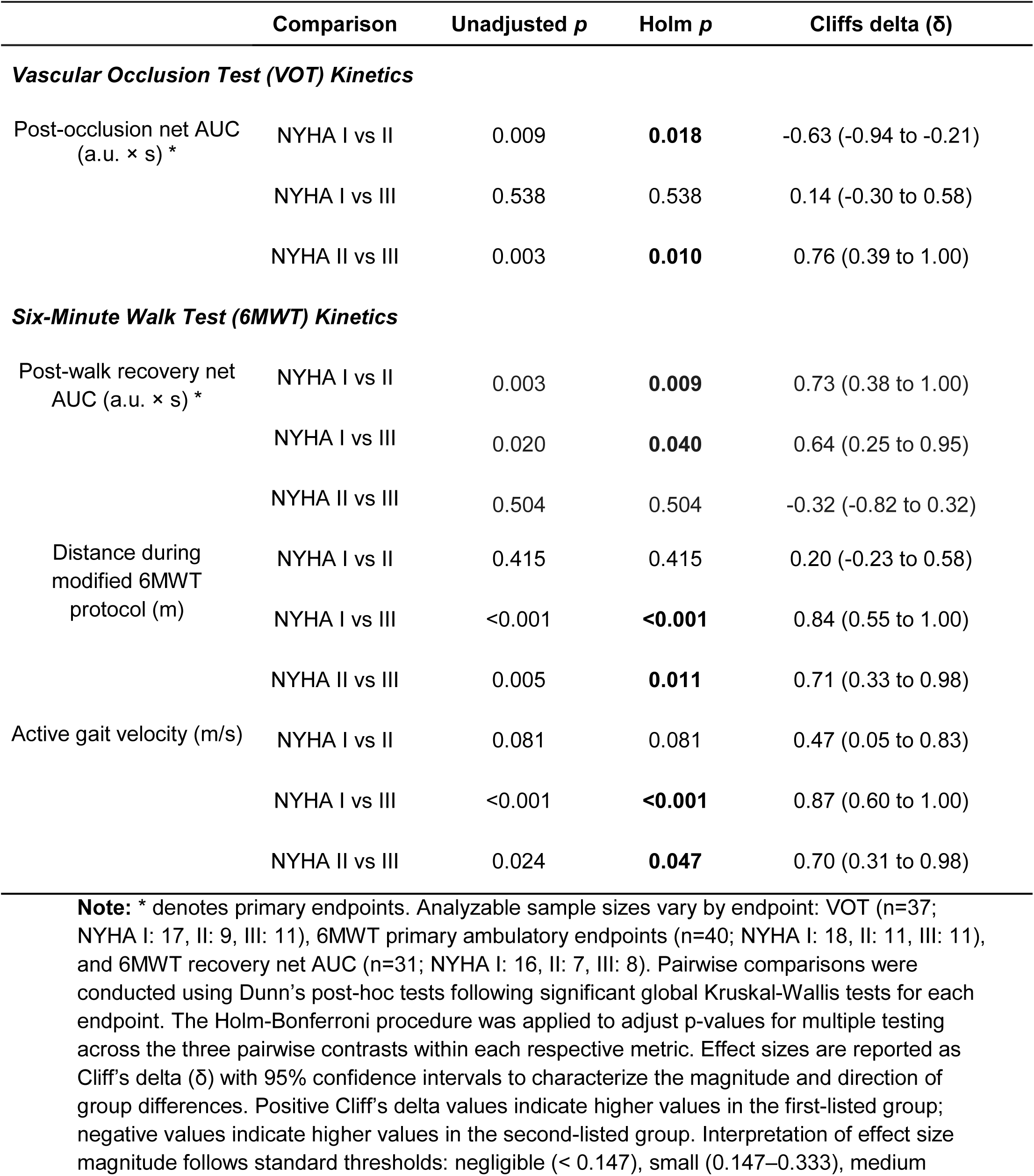
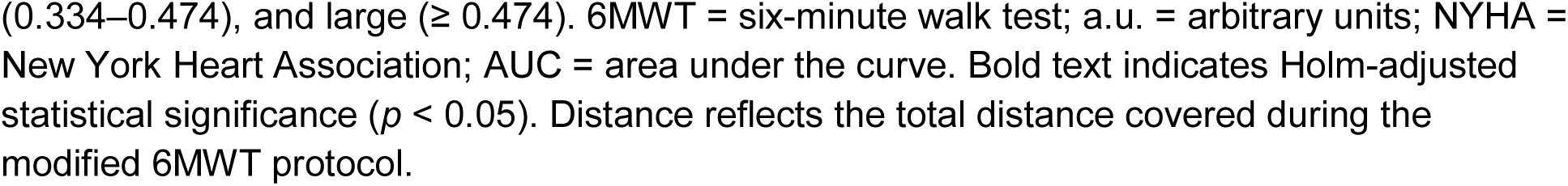
Pairwise post-hoc comparisons of significant physiological and functional endpoints by NYHA class.

#### Post-walk recovery netAUC across New York Heart Association functional classification

Post-walk recovery net AUC differed across classes (H = 10.95, p = 0.004, ε² = 0.32; Figure 4). NYHA I showed a net reoxygenation overshoot (median 46.1 a.u.×s), whereas NYHA II (−16.3) and NYHA III (−12.8) showed net oxygen deficits; NYHA I differed significantly from both II and III, with no difference between II and III (Table 3). Findings were consistent in the zero-clipped pAUC sensitivity analysis.

### Secondary vascular occlusion test analyses

Among secondary VOT endpoints, the maximum reperfusion slope did not reach significance (H = 5.78, p = 0.056) but was descriptively steepest in NYHA II, and the reactivity ratio did not differ across classes (p = 0.338; Table 2). Ischemic-stimulus metrics (occlusion duration, ischemic nadir, maximum deoxygenation slope) were comparable across classes (all p ≥ 0.252; Supplementary Table S3.

### Secondary six-minute walk test analyses

Total distance during the modified 6MWT differed across classes (p < 0.001): NYHA III walked less (280 m) than NYHA I and II (both 420 m; Table 3). Active gait velocity showed the same gradient (NYHA I, 1.19; II, 1.02; III, 0.74 m/s; p < 0.001). The exertional Oxy nadir did not differ across classes (p = 0.722). Total elapsed test time and rest-break frequency both increased with class (p < 0.001 and p = 0.007, respectively).

#### Exploratory correlations between VOT and 6MWT endpoints

In exploratory correlations (n = 30–33; Figure 5), the VOT reactivity ratio was positively associated with gait velocity (r_s_ = 0.42) and walk distance (r_s_ = 0.46), and the maximum reperfusion slope with walk distance (r_s_ = 0.41). The post-walk recovery net AUC correlated with both gait velocity (r_s_ = 0.44) and walk distance (r_s_ = 0.39). The post-occlusion net AUC and hyperemic pAUC showed no significant functional associations, and the VOT and 6MWT area-based endpoints were uncorrelated with one another (all |rs| ≤ 0.28).

**Figure 5.**
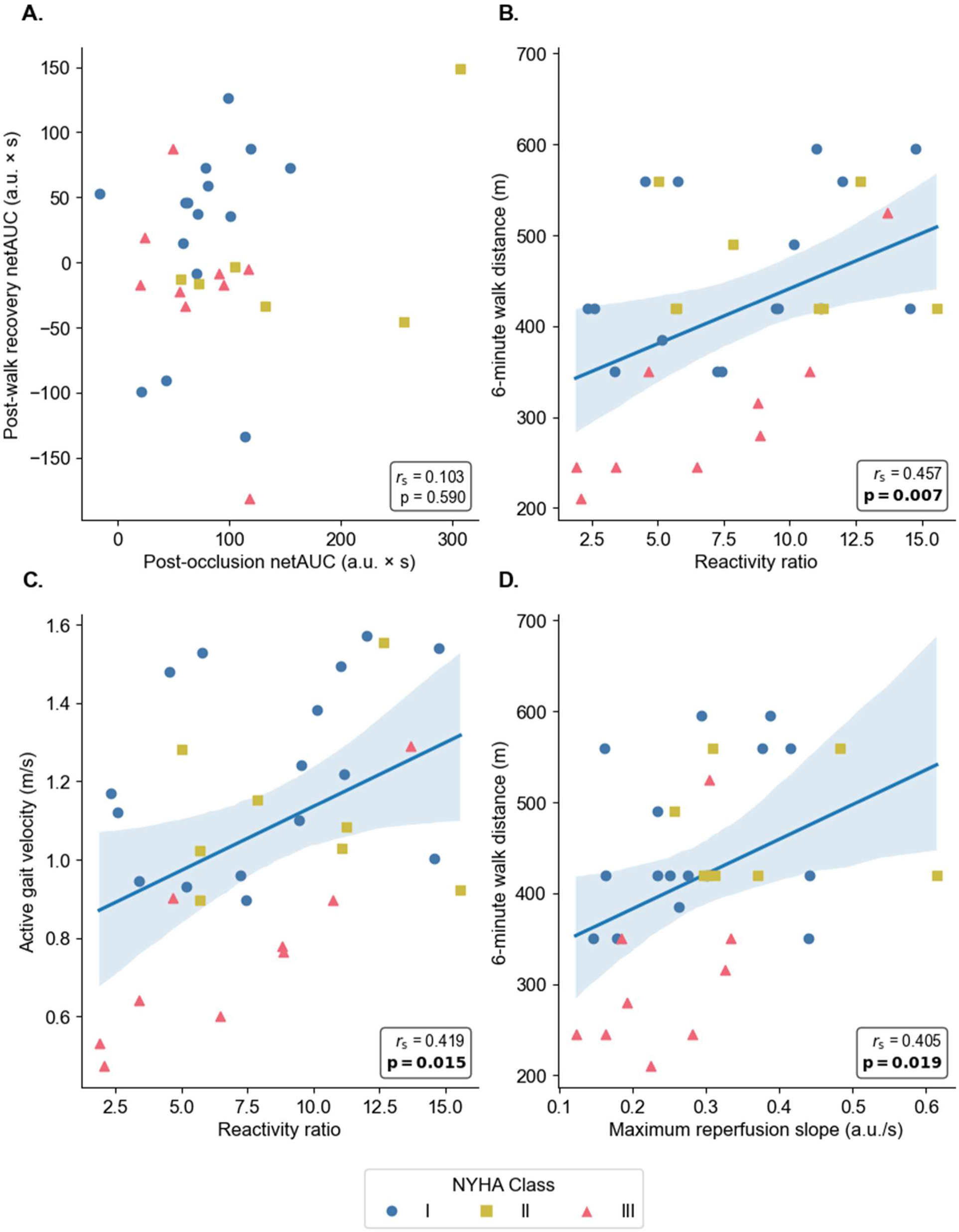
Associations between resting microvascular reserve and functional ambulatory performance. *Note*. Scatterplots illustrating exploratory Spearman rank-order correlations (r_s_) between paired resting vascular occlusion test (VOT) and 6-minute walk test (6MWT) endpoints (n = 33). (A) Post-occlusion net area under the curve (net AUC) versus post-walk recovery net AUC. (B) Reactivity ratio versus 6-minute walk distance. (C) Reactivity ratio versus active gait velocity. (D) Maximum reperfusion slope versus 6-minute walk distance. Solid blue lines represent the linear line of best fit, with shaded regions denoting 95% confidence intervals; regression lines are only displayed for panels where the overall association reached statistical significance (*p* < 0.05). Net AUC values represent the algebraic (unclipped) baseline-referenced integral and therefore include values falling below the resting baseline. Individual data points are shaped and color-coded by New York Heart Association (NYHA) functional class (I: blue circles; II: yellow squares; III: red triangles). Spearman coefficients (r_s_) and *p*-values are displayed within each panel, with statistically significant findings (*p* < 0.05) bolded. a.u. = arbitrary units; AUC = area under the curve; 6MWT = six-minute walk test; NYHA = New York Heart Association; VOT = vascular occlusion test. Distance reflects the total distance covered during the modified 6MWT protocol.

#### Device Tolerability and Patient Acceptability

Most participants rated the device as comfortable or very comfortable at the sternal (93%) and forearm (95%) sites, and the adhesive as comfortable on the chest (91%) and forearm (93%). Most (91%) anticipated that self-application would be easy or extremely easy. No participant reported a maximum wear time under one day, and half (50%) were willing to wear the chest sensor for more than 7 consecutive days (forearm, 47%).

## Discussion

This pilot study evaluated a novel wearable NIRS-based tissue oximetry system (NIRSense Envello Core) during resting (VOT) and ambulatory (6MWT) physiological stressors in chronic HF outpatients. Post-walk recovery net AUC differentiated NYHA I from NYHA II, despite these classes recording identical distances during the modified 6MWT protocol. Resting VOT profiling revealed an unexpected non-monotonic pattern, with the greatest post-occlusion net AUC and reactive hyperemia pAUC observed in NYHA II. The novel wearable device demonstrated excellent patient acceptability and tolerability. Collectively, these preliminary findings suggest that NIRS-augmented testing during physiological stressors offers a promising, non-invasive framework for objective physiological phenotyping in outpatient heart failure. Given its simplicity, this could also be used to track patients’ performance over time, potentially without the 6MWT at times.

### 6MWT

A novel aspect of this study was the use of a wearable NIRS platform to continuously monitor tissue oxygenation during and immediately after the 6MWT, an approach previously unfeasible due to the restrictive form factor and tethering of conventional systems. Post-walk net AUC differed between NYHA I and NYHA II, despite these classes recording identical median distances during the modified 6MWT. This is in line with other research showing that traditional outcomes like total distance can fail to differentiate early chronic HF disease stages^6^. This divergence between tissue-oxygenation recovery and ambulatory distance was not confined to the NYHA-stratified analysis. In classification-independent exploratory correlations, the post-walk recovery net AUC was positively and moderately associated with both gait velocity and 6-minute walk distance, indicating that the integrated oxygenation response scales with functional capacity across the continuous performance spectrum rather than only at discrete and subjective NYHA boundaries. Our findings should be interpreted cautiously given the pilot sample size but may suggest that post-exertional tissue oxygenation recovery provides unique physiological information beyond traditional clinical metrics. This highlights the potential of NIRS-derived recovery kinetics as objective markers to detect early functional impairment during the NYHA I-to-II transition, before overt reductions in ambulatory distance manifest.

There were no significant differences in exertional Oxy nadir across NYHA class. One plausible explanation is that patients regulate walking intensity through behavioral modifications such as self-pacing, and this is supported by our velocity data, such that NYHA III participants walked 27% and 38% slower than NYHA II and NYHA I participants, respectively. Moreover, total elapsed test time increased across NYHA classes despite the fixed 360-second active locomotion period, with NYHA II and III requiring longer elapsed time than NYHA I. Finally, rest-break frequency also differed across classes, driven by more breaks in NYHA III versus NYHA I. However, because perceived exertion, heart-rate response, and other objective effort markers were not available, the contribution of self-pacing cannot be directly confirmed in the present study.

### VOT

An unexpected non-monotonic hyperemic pattern emerged in this study, whereby VOT reactive hyperemia pAUC and post-occlusion net AUC were greatest in NYHA II. This contrasts with a prior report in which forearm microvascular reactivity was reduced in chronic HF but did not differ across disease severity^25^. The enhanced reactive hyperemia observed in NYHA II may instead reflect a compensatory increase in microvascular reserve at an intermediate disease stage, whereas the lower net AUC and pAUC in NYHA III may indicate attenuation of this reserve. Consistent with such attenuation, worsening functional class carries an increasing risk of sarcopenia and frailty, which have been shown to reduce VOT-derived vascular responsiveness and blunt post-occlusive reperfusion kinetics in older outpatients with chronic HF^26^

The hyper-reactive AUCs observed in NYHA II were not accompanied by statistically significant differences in occlusion duration, ischemic nadir, or maximum deoxygenation slope, suggesting that the pattern was unlikely to be explained solely by a larger ischemic stimulus. Consequently, this finding more likely reflects microvascular reserve rather than an artifact of varying ischemic exposure. Nevertheless, residual confounding remains possible. Medication use differed across NYHA classes, and the pilot sample size precluded multivariable adjustment. For example, beta-blocker prescription was notably lower in NYHA II (75%) than in NYHA I (95%) and NYHA III (92%), a difference whose effect on hyperemic responses is uncertain^27^. SGLT2 inhibitor and diuretic use increased progressively from NYHA I to III (42% to 69% and 11% to 54%, respectively), and SGLT2 inhibitors have been shown to modulate peripheral microvascular function in HF^28^. MRA prescription also exhibited a non-monotonic distribution (NYHA I: 58%, II: 25%, III: 69%) that partially mirrors the patterns in net AUC and pAUC. Future studies should incorporate dose-level pharmacological data and dose-normalized hyperemic responses to disentangle whether this non-monotonic pattern reflects intrinsic compensatory vascular reserve, treatment effects, or residual confounding.

### Cross-stressor analysis

The VOT and 6MWT metrics appeared to provide non-redundant physiological information, resolving different regions of the disease-severity spectrum. The post-walk recovery net AUC differentiated NYHA I from both NYHA II and NYHA III but did not separate NYHA II from NYHA III, indicating particular sensitivity to early functional impairment. In contrast, the VOT net AUC and modified 6MWT distance distinguished the more advanced NYHA II-to-III stages. Consistent with this complementarity, the VOT and 6MWT area-based endpoints were not correlated. Together, these findings suggest that resting ischemia–reperfusion responses and post-exertional recovery kinetics capture related but non-interchangeable components of the HF physiological phenotype. This supports the rationale for a paired-stressor NIRS approach and emphasizes the need for larger longitudinal studies to determine which features provide prognostic or treatment-monitoring value.

Additionally, resting VOT-derived reperfusion kinetics were positively associated with 6MWT performance: the reactivity ratio correlated with both 6-minute walk distance and active gait velocity, and the maximum reperfusion slope correlated with walk distance, though not gait velocity. These associations indirectly suggest that intrinsic peripheral vascular reactivity contributes to the limitation of functional capacity in chronic HF. From a practical standpoint, the 6MWT is time-consuming, space- and staff-intensive^21^, and dependent on patient effort and ambulatory ability, which limits its feasibility for routine or serial assessment. A brief, passive, resting microvascular test that tracks ambulatory capacity could therefore offer a more scalable and effort-independent surrogate, potentially extending functional assessment to patients unable to complete a walk test. However, these exploratory associations should be interpreted cautiously given the small sample size and potential class-driven effects

### Potential translational implications

Our data supports the feasibility of embedding wearable tissue oximetry into outpatient functional testing to identify physiological abnormalities not captured by traditional metrics. Moreover, the untethered, miniaturized sensor evaluated in this study demonstrated excellent patient acceptability. Over 90% of participants reported high comfort scores for both the device and its biocompatible adhesive, alongside high anticipated ease of self-application. Participants also expressed a strong willingness to adopt the sensor for multi-day monitoring, with half the cohort amenable to consecutive wear periods exceeding one week. Together, these findings indicate that integrating NIRS-based microvascular phenotyping into routine outpatient care, or even extending it to home-based monitoring, may be both methodologically feasible and highly tolerated by patients with chronic HF.

### Methodological considerations

Several limitations should be acknowledged. First, this pragmatic single-center pilot had a modest sample, and endpoint-specific signal loss reduced the analytic sample, particularly for post-walk recovery net AUC; subgroup-versus-cohort comparisons were underpowered and cannot exclude selection bias. Second, because the timer was paused during rest breaks, total 6MWT distance is not comparable with ATS norms and is likely higher. Third, the sternal NIRS signal reflects a heterogeneous tissue domain and should be read as a mixed systemic-regional recovery signal, not an isolated locomotor-muscle measure^23^; adipose thickness can influence amplitude at the 35-mm separation used^22,29^, and wider separations were avoided owing to attenuated light and motion susceptibility^30^. Fourth, the absence of LVEF, natriuretic peptides, and objective effort markers limits mechanistic interpretation and precludes comparison across HF phenotypes. Fifth, medication use differed across classes, and the sample size precluded multivariable adjustment. Sixth, without a healthy age-matched control, NYHA I serves as a relative rather than normative comparator. Seventh, the tolerability questionnaire was an ad hoc, non-validated instrument. Finally, the cross-sectional design precludes causal inference; longitudinal studies are needed to establish whether these NIRS features predict deterioration, treatment response, or HF hospitalization.

Important strengths include the paired stressor paradigm, which enabled evaluation of both resting ischemia-reperfusion responses and post-exertional recovery kinetics during a single outpatient visit. The fully untethered and portable sensor enabled the continuous characterization of the previously unexplored post-walk recovery window. Additionally, the cohort included broad Fitzpatrick skin type representation (25% types V-VI), supporting preliminary feasibility across diverse skin pigmentation levels. Finally, test-retest reliability was formally assessed for primary VOT endpoints, providing initial evidence that wearable NIRS-derived reactive hyperemia metrics are sufficiently reproducible for between-group comparisons.

### Future research directions

Future multi-center longitudinal studies are warranted to validate our findings and their prognostic value for clinical endpoints, including heart failure hospitalization and mortality. Investigations should also expand across pathophysiological phenotypes (HFrEF versus HFpEF) to capture distinct microvascular mechanisms. Larger cohorts will also enable the use of multivariable modeling or propensity-score matching to control for baseline pharmacological differences and ensure that observed microvascular variations are isolated to disease severity rather than medication effects. Prospective randomized trials should evaluate whether NIRS-augmented assessments, particularly 6MWTs pre- and post-GDMT initiation, can track dynamic therapeutic responses to medical management, mechanical circulatory support, and other heart failure therapies. If found to be the case, this non-invasive approach could accelerate GDMT titration, potentially reduce the overarching cost of care, and act as a biofeedback tool to improve patient medication adherence. Finally, extending wearable NIRS applications to simultaneously monitor cerebral tissue oxygenation offers a critical next step. While continuous exertional cerebral hemodynamics have recently been characterized in healthy cohorts^31^, evaluating concurrent cerebral hypoperfusion in HF could unmask distinct central limitations to exercise tolerance that complement the peripheral recovery kinetics observed in this study.

## Conclusion

In this single-center pilot study of 44 chronic HF outpatients, a wearable tissue oximeter (NIRSense Envello Core) revealed distinct oxygenation patterns across NYHA classes. Post-walk recovery net AUC differed between NYHA I and II despite identical distance-based performance, suggesting NIRS recovery kinetics capture physiological information inaccessible to functional capacity metrics. Resting VOT profiling showed an unexpected non-monotonic hyperemic response, greatest in NYHA II, which may reflect compensatory vascular reserve or residual medication confounding. The device was well tolerated, with over 90% reporting high comfort and willingness for extended home monitoring. These findings establish proof-of-concept for integrating wearable NIRS-augmented stressor testing into outpatient HF evaluation and warrant validation in larger, longitudinal cohorts with prognostic endpoints and comprehensive HF phenotyping.

## Data Availability

The data that support the findings of this study are available from the corresponding author upon reasonable request.

## Acknowledgements

The authors acknowledge the patients who generously volunteered their time and effort to participate in this study. We also wish to recognize Rachel Junkin and the entire staff at Jackson Heart Clinic for their outstanding clinical coordination and operational support throughout the research process.

